# Towards Participatory Precision Health With Co-designed Recommendations for Just-in-Time Adaptive Interventions in Adolescents and Young Adults: A Systematic Review

**DOI:** 10.1101/2025.09.01.25334834

**Authors:** Kathleen W. Guan, Mohammed Amara, Sıla Gürbüz, Imran Khan, Christopher Adlung, Viviana Cortiana, Rayyan Ali, Carmine Iorio, Eva Thalassinou, Crystal R. Smit, Annabel Vreeker, Eeske van Roekel, Loes Keijsers, Mark de Reuver, Caroline A. Figueroa

**Affiliations:** Faculty of Technology, Policy, and Management, Delft University of Technology, The Netherlands; Department of Experimental Psychology, University of Oxford, United Kingdom; Faculty of Medicine, Karadeniz Technical University, Turkey; Arkansas College of Osteopathic Medicine, United States; Department of Medical and Surgical Sciences, University of Bologna, Italy; Imperial College School of Medicine, United Kingdom; Department of Philosophy, Social Sciences and Education, University of Perugia, Italy; Department of Research and Development, Gro-up, The Netherlands; Erasmus School of Social and Behavioural Sciences, Erasmus University Rotterdam, The Netherlands; Department of Child and Adolescent Psychiatry and Psychology, Erasmus University Medical Center, The Netherlands; Department of Developmental Psychology, Tilburg University, The Netherlands

**Author notes:** Corresponding author: Kathleen W. Guan.

**Keywords:** Adolescents, young adults, just-in-time adaptive interventions, mobile health, health behaviors, precision health

## Abstract

**Background:** The transition from adolescence to young adulthood (10-25 years) constitutes a sensitive developmental period marked by rapid biological, psychological, and social change, during which preventive health interventions can shape long-term outcomes. Mobile health (mHealth) tools offer accessible opportunities for tailored support for this population, but often adapt poorly to dynamic contexts, resulting in inconsistent engagement and effects. Just-in-time adaptive interventions (JITAIs), which tailor support in real time by leveraging ongoing data, are increasingly explored as precision health strategies. However, how these mechanisms are designed, implemented, and evaluated for adolescents and young adults (AYAs) has not yet been systematically reviewed.

**Objectives:** This review aimed to synthesize the evidence on JITAIs developed for AYAs, examine how their adaptive mechanisms have been designed to support specific health goals and changing AYA contexts, and assess methodological quality of reporting to inform future precision health intervention development.

**Methods:** We conducted a systematic review in accordance with Preferred Reporting Items for Systematic Reviews and Meta-Analyses (PRISMA) and Synthesis Without Meta-analysis (SWiM) reporting guidelines. Twelve databases were searched for peer-reviewed studies published from 2013 to 2025. Eligible studies were peer-reviewed, focused on participants aged 10 to 25 years, and reported real-time adaptive mobile health interventions consistent with JITAI design principles. Two reviewers independently conducted screening, data extraction, and methodological quality appraisal using Joanna Briggs Institute checklists. AYA co-authors contributed to all phases of the review. Due to substantial heterogeneity in study populations, intervention content, adaptive mechanisms, comparators, and outcome measurements, findings were synthesized narratively and no meta-analysis was conducted.

**Results:** 61 unique interventions were included. JITAIs for AYAs addressed substance use (N=24, 39.3%), mental health (N=23, 37.7%), and physical health or chronic conditions (N=14, 23%). JITAI tailoring mechanisms relied predominantly on self-reported behavioral data. Decision rules were typically symptom threshold-based and decision points were commonly daily or event-triggered. Methodological concerns with reporting on intervention administration, participant selection, and outcome measurement reliability were pervasive across all studies, limiting interpretability of observed effects and cross-study comparisons. Ethical considerations, including researcher positioning and reflexivity, alongside the depth of reporting around participatory AYA engagement in design and implementation, were also inconsistent.

**Conclusion:** This review contributes a novel perspective to AYA digital health by moving beyond intervention outcomes to systematically examine how core adaptive mechanisms are operationalized for AYAs across multiple health domains, while also directly integrating AYA perspectives into the interpretation of findings and recommendations for future work. In contrast to prior reviews focused primarily on adults or specific conditions, it identifies broader contextual, methodological, and ethical considerations relevant to AYA precision health. Taken together, our findings highlight the critical need for more transparent, contextually responsive, and youth-centered adaptive interventions, alongside more rigorous designs for evaluating adaptive intervention components in daily life contexts.

PROSPERO registration: CRD42023473117

## INTRODUCTION

Adolescence is a formative period for preventative health interventions. As they transition into young adulthood, adolescents experience continued brain and physical development, alongside heightened vulnerability to risk factors for poor health [1,2]. For example, partaking in behaviours that compromise health, such as substance use, poor diet, and insufficient physical activity, in early life can increase the risk of chronic diseases such as cancer [3], and disrupt key neurodevelopmental processes related to stress regulation and executive functioning [4]. Further, the transition to adulthood is a complex developmental stage, shaped by rapidly shifting and interrelated biological, psychological, and social changes that are unique to each individual [5]. Behavioral patterns during this period emerge dynamically in response to fluctuating contexts and interactions, with significant implications for long-term health. For example, food insecurity may predispose adolescents to poor dietary habits, which in turn can affect their physical health, cognitive development, and emotional resilience over time [6]. In light of these vulnerabilities, early and precisely targeted interventions during this period are critical for mitigating both immediate and long-term health risks to promote healthier developmental pathways. Given the record number of adolescents and young adults (AYAs, ages 10 to 25 years) globally [7], investing in tailored health interventions during adolescence can significantly improve population health outcomes while alleviating pressures on health systems [8].

Precision health represents a significant paradigm shift from generalized medical approaches, by prioritizing adaptive strategies that tailor health support to optimize diverse outcomes unique to each individual [9]. Whilst much of the initial application of precision health has occurred within adult medical contexts [8], there is growing recognition that such approaches may be particularly suited to addressing the multifaceted challenges during the transition to adulthood [10]. Importantly, precision health approaches during this unique period can enhance both the relevance and effectiveness of AYA health interventions. In particular, addressing behaviours and states calls for interventions that precisely tailor to individual risk factors and contextual influences shaping AYAs’ lives.

Mobile health (mHealth) technologies are a key avenue for delivering tailored health support to AYAs, whose pervasive use of digital platforms throughout their daily lives and affinity for mobile communication provide opportunities for accessible intervention delivery. Such tools have demonstrated some potential to support and promote positive health behaviours, such as increasing physical activity, improving nutrition, and reducing substance use among adolescents [11]. However, mHealth interventions often produce inconsistent effects, due to challenges in delivering engaging content that is precisely relevant to AYAs’ evolving health contexts over time [12,13]. Extant literature has suggested that use of digital tools to support health behaviours may decline when interventions fail to resonate with or adapt to changing circumstances [14,15]. Demographics (e.g., age, gender), psychosocial characteristics (e.g., mood), and contextual factors (e.g., setting of delivery) play a significant role in individual engagement with mHealth interventions [13]. Thus it is necessary to identify key considerations for designing mobile interventions that are not only tailored, but also dynamically and temporally responsive to AYAs’ shifting needs.

One particularly promising precision health approach that has emerged is the just-in-time adaptive intervention (JITAI). JITAIs utilize ongoing data from mobile sensing technology with the aim to deliver tailored contextually relevant support precisely when it is most needed for each individual [16]. JITAIs can provide a flexible strategy for promoting positive health behaviours across individually salient moments and contexts by dynamically adjusting content in response to individual changes. The temporal adaptability that characterizes JITAIs is particularly important given the variability in AYA development, as their health risks and support needs can rapidly shift based on environmental and intrapersonal factors such as mood, stress levels, and social interactions from moment to moment [17,18]. JITAIs can crucially enhance the responsiveness of precision health strategies by delivering support attuned to ongoing changes in AYA contexts, particularly when they can be receptive to this support, which can help ensure tailored intervention content remains relevant and effective in addressing evolving needs.

Despite their potential, most prior analyses of JITAIs have focused on older adults or specific health conditions [19–24]. As such, there is a limited understanding of how JITAIs can support the dynamic health needs of AYAs as they transition into early adulthood. Given the rapidly changing and diverse contexts that characterize this formative developmental stage, understanding how JITAIs currently adapt to this period (10 to 25 years) is critical.

This review thus had three specific objectives. First, it aimed to map and synthesize the evidence base on JITAIs developed for AYA health contexts. Second, it aimed to characterize how key adaptive components—namely tailoring variables, intervention options, decision rules, and decision points [16]—were designed and applied in relation to targeted AYA health outcomes. Third, it aimed to critically appraise the quality of the included studies in order to identify methodological and evidence gaps. Overall, the findings are intended to inform the design of more effective, personalized, and contextually responsive mobile health interventions to support healthier trajectories from adolescence into adulthood and beyond.

## METHODS

This systematic review followed reporting guidelines from Preferred Reporting Items for Systematic Reviews and Meta-Analyses (PRISMA) and SWiM (Synthesis Without Meta-analysis), respectively [25,26]. The review was pre-registered in the PROSPERO database under registration number CRD42023473117. The review protocol was also published after peer review [27].

The search strategy for the review was developed in consultation with an information specialist, and rationale for the search strategy, including scope of dates and search terms, is reported in the supplementary material according to PRISMA-S [28]. The search strategy employed a combination of keywords and database-specific subject headings (e.g., MeSH terms) to capture peer-reviewed literature related to JITAIs, AYAs, digital health technologies, and health outcomes and was peer-reviewed [27]. The literature search took place across multiple electronic databases to identify relevant literature from 2013 to 2025; these databases included PubMed, Scopus, Web of Science, PsycINFO, Embase, Cumulative Index to Nursing and Allied Health Literature, Institute of Electrical and Electronics Engineers Xplore, Association for Computing Machinery Digital Library, Cochrane Central, Dimensions, World Health Organization Global Index Medicus, and Google Scholar.

The individual database searches were conducted in July 2025, with additional keyword-based searching in Google Scholar until December 2025. Reference list screening of extracted studies was used to identify further eligible studies. Further, studies included in prior systematic reviews of JITAIs were consulted [19–24], which enabled inclusion of all relevant studies published prior (as the literature search strategy only started in 2013, the first explicit mention of JITAI and AYAs in the literature [29,30]). These steps helped ensure comprehensive coverage of past and emerging studies. It was not deemed necessary to contact any study authors.

### Screening, inclusion/exclusion, and extraction procedure

All literature search records were imported into Covidence, a systematic review text management platform, with duplicates automatically removed. Each title and its accompanying abstract was screened by two reviewers (either KWG, CA, CI, IK, and/or RA each independently), following a pilot screening phase to ensure consistency in eligibility decisions. Full texts were also screened by two reviewers each. All discrepancies during screening or assessment of full texts were resolved through discussion. Study inclusion was primarily determined by whether studies provided sufficient detail on JITAI components to inform the review’s focus on how these interventions adapt to AYA contexts and health needs. Studies were eligible for inclusion if they were peer-reviewed, focused on AYAs aged 10 to 25 years, and reported mHealth interventions that adapted in real time to user data, consistent with the defining features of JITAIs. Other mHealth studies were out of scope, including JITAI studies that involved participants beyond the AYA age range (determined by reported sample age range and mean age, where available) and did not report these findings separately [31]. We selected the age range of 10 to 25 years, given its increasing recognition as its own heterogeneous yet distinct developmental period among scholars [10,32–34]. We did not restrict by study type or design, such as control conditions. We also excluded JIT simulation studies that did not involve evaluating an intervention [35].

Data from each study were also extracted by two reviewers (either KWG, SG, VC, and/or RA each independently) using a piloted, standardized data extraction form to ensure consistency. The data extraction form followed standardized conceptualizations of JITAI in the literature [16,27], which outlines key components guiding interventions to be adapted to individual needs: intervention options, or the different forms of support or methods for delivering that health support may be considered based on tailoring variables; tailoring variables, referring to the data collected about an individual, which informs how and when an intervention should be adapted; and finally, decision rules, or the guidelines that use these tailoring variables to determine the selection, timing, and delivery of intervention options at specific decision points or specific moments, such as particular times of the day or contextual triggers when these decisions are made [16,27]. These components together shape how JITAIs dynamically adjust to ensure interventions remain relevant across shifting adolescent contexts [16]. By structuring the form around these components, the method was aligned with the review’s focus on mechanisms of adaptability in JITAIs.

Further, distal and proximal outcomes were also extracted, as they indicate the health objectives toward which tailoring mechanisms are directed. Specifically, distal outcomes represent the overarching, long-term goals of a JITAI, while proximal outcomes are short-term health objectives that often serve as mediators that lead towards these distal targets [16]. Thus, data extracted from each study included participant characteristics, intervention details, tailoring mechanisms, study design, and outcomes reflecting the goals of tailoring mechanisms (both proximal and distal). See supplementary materials for the full extraction data for each study.

No formal amendments were made to the registered or published protocol [27]. Several additional outcomes of interest, including user engagement and ethical considerations, were identified in the protocol as potentially informative for interpreting effectiveness; however, inconsistent measurement and incomplete reporting meant they could not be systematically synthesized or graded. While information on AYA engagement and ethical considerations was documented during data extraction for each study where reported, reporting was sparse and also lacked sufficient detail for structured synthesis.

### Risk of bias and justification for synthesis procedure

Two reviewers (KWG, MA) independently assessed methodological quality, including risk of bias, using the Joanna Briggs Institute (JBI) checklists for distinct study designs, with discrepancies resolved through discussion [36]. The JBI checklists for each study included critical appraisal according to temporal precedence, selection and allocation, confounding factors, intervention administration, outcome measurement (timepoints, method, and reliability) participant retention, and statistical conclusion validity, which we report in the results section.

These appraisals informed consideration of the Grading of Recommendations Assessment, Development, and Evaluation (GRADE) framework [37], which was prespecified in the protocol to assess certainty of evidence across studies. However, application of GRADE across studies requires sufficient comparability in outcome definitions, measures, and reporting [38], and this was not present within any outcome domain in the current review. We therefore did not undertake a formal assessment of evidence certainty or reporting bias. Further, although studies could be grouped broadly by outcome domain, substantial heterogeneity remained in participant populations and settings, intervention content and delivery, tailoring mechanisms, comparator conditions, and outcome assessment. We therefore also did not conduct a meta-analysis because we did not deem the included studies sufficiently similar to support a meaningful pooled estimate. Findings were instead synthesized narratively and organized according to key JITAI tailoring components and outcome domain, including intervention aims, tailoring variables, decision rules, and decision points. All included studies were considered in the narrative synthesis.

### AYA engagement procedure

To ground implications of this review in lived experience, we directly engaged 12 AYAs, including health activists from the World Health Organization European Region’s Youth4Health network, a platform for AYA participation in health policymaking. These AYAs, including young entrepreneurs, medical students, and policy advocates, provided critical perspectives that informed study interpretations and implications. Seven were directly involved as co-authors in screening studies, extracting data, and conducting critical appraisals, including the lead author. AYA perspectives were further elicited through a dedicated virtual consultative session with AYAs who were not part of the co-authorship team. During this session, five AYAs reviewed the data extraction tables from the review and engaged in semi-structured critical dialogue regarding intervention acceptability, design relevance, and the alignment of findings with lived adolescent experiences. These discussions were documented via meeting minutes and synthesized by KWG and MA to identify recurring interpretive themes. Given several of the AYA participants’ role as co-authors in the review alongside the limited sample, these AYA contributions are reported in the discussion section as explicitly interpretive observations to contextualize preliminary recommendations for future work, rather than as additional empirical data derived from the included studies.

## RESULTS

Following this exclusion, a total of 252 full text articles were assessed for relevance. Studies that were excluded at the full text stage did not develop mobile interventions that were tailored to dynamic adolescent contexts, for example, those offering only pre-scheduled digital health support without ongoing adaptation; therefore not meeting the definition of a JITAI [16]. This resulted in a final total of 69 studies (with 61 unique interventions) developing JITAIs for AYAs included in the findings synthesis that follows.

**Figure 1.**
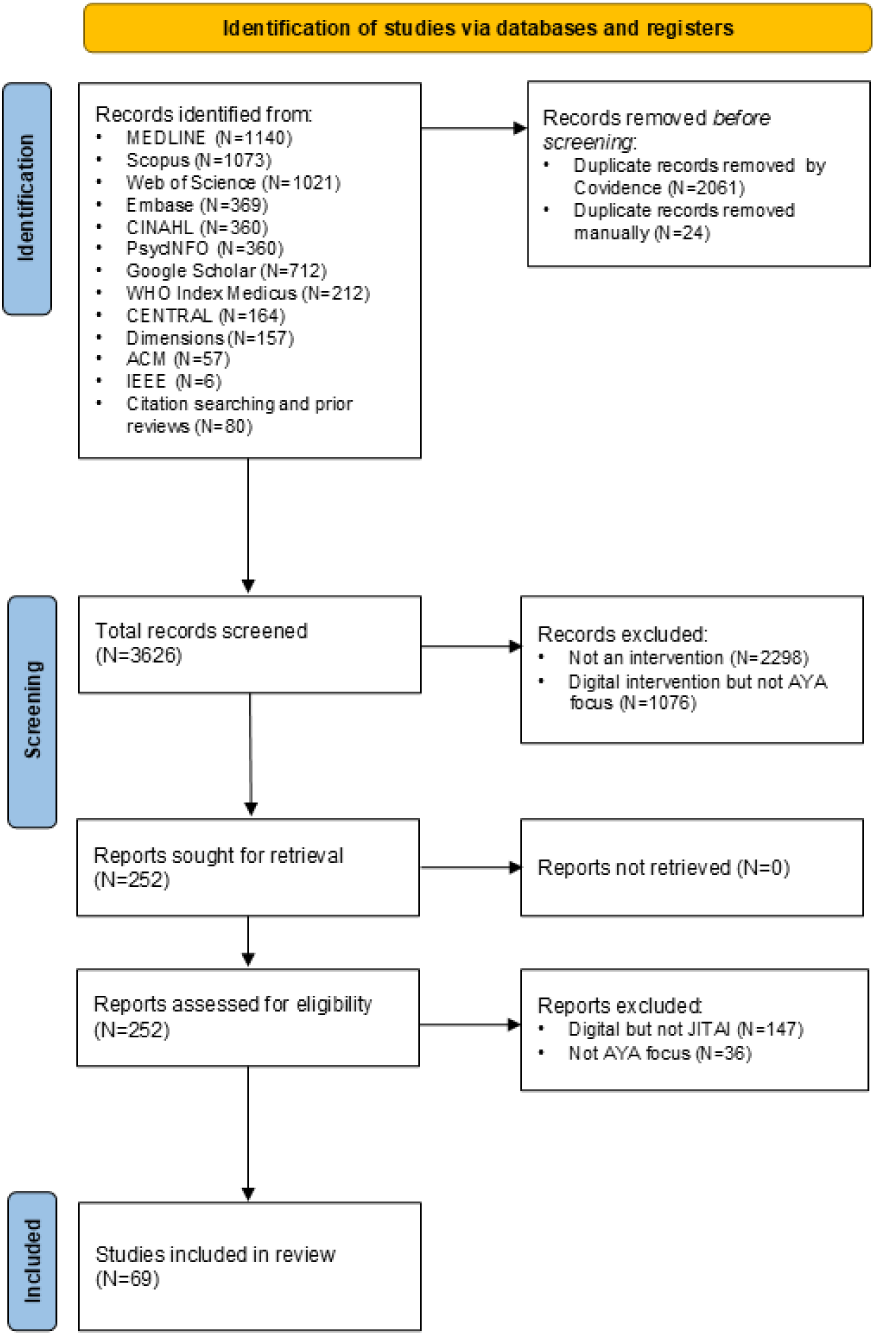
PRISMA flow diagram for AYA JITAI study selection 3,626 unique records were identified via literature search and screened in the title/abstract phase (Figure 1). 3,374 records were excluded as they were clearly with adult populations or not intervention studies, such as studies collecting ecological momentary assessments only.

### Risk of bias and quality of evidence

The risk of bias and critical appraisal matrix (Figure 2) shows that every study had at least one methodological concern, indicating moderate to high risk of bias present across the evidence base. We organize these by study design (randomized controlled trial or RCT, non-controlled experimental studies, and qualitative studies) as critical appraisal is conducted based on the specific design of a study [36]. Importantly, many “unclear” ratings are likely attributable to incomplete reporting rather than the absence of methodological safeguards, reflecting variability in reporting practices and the lack of standardized conventions in this field. These issues significantly reduce confidence in reports of effectiveness and complicate comparisons across studies.

**Figure 2:**
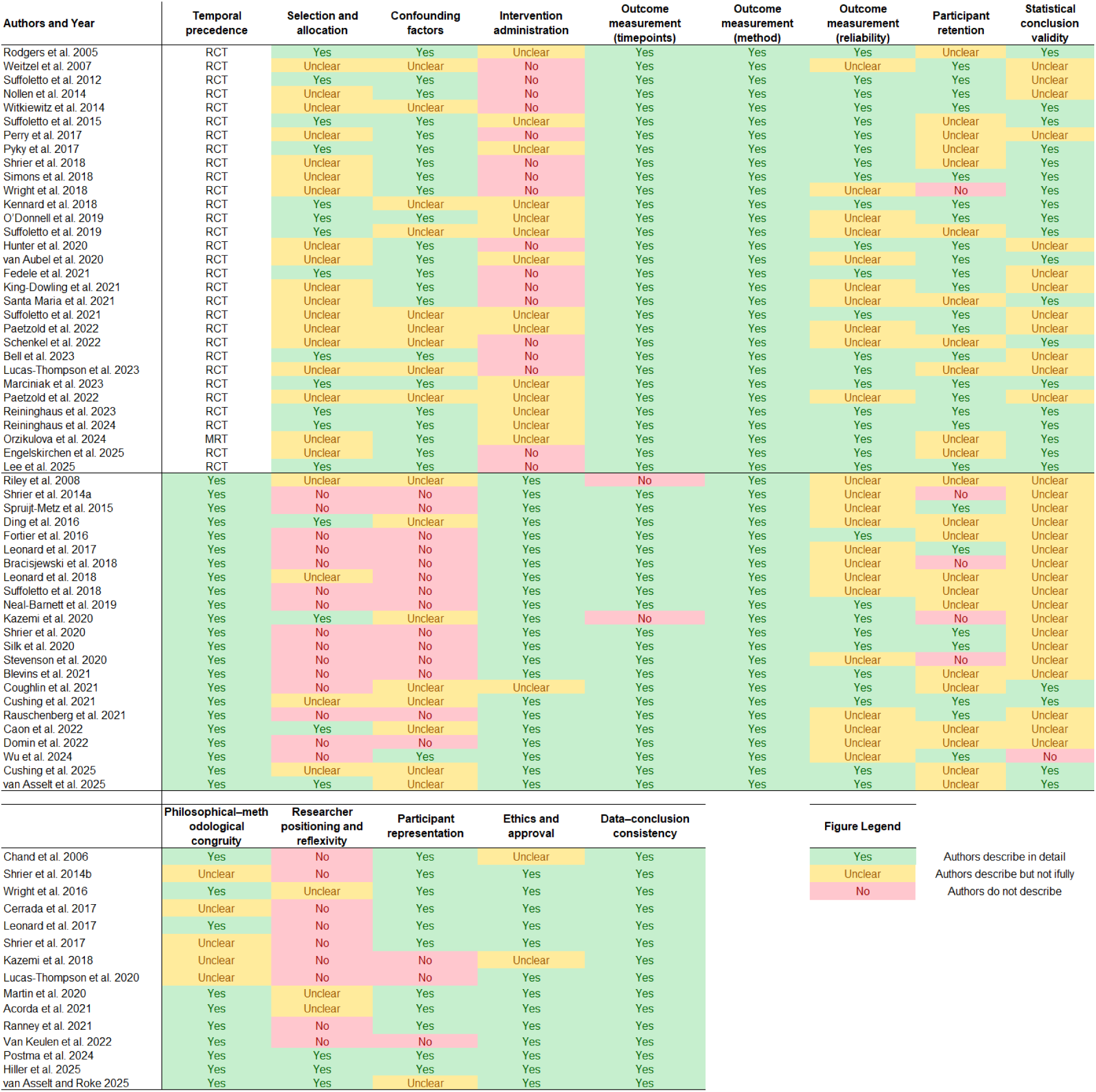
Risk of bias and methodological quality appraisal of randomized controlled trials, non-controlled experimental studies, and qualitative studies of AYA JITAIs

Intervention administration was the most prominent limitation unique to RCTs. Across studies, reporting was limited on whether treatment groups were treated identically apart from the intervention of interest. Importantly, intervention administration is inherently difficult to deliver in a blinded and standardized manner in JITAIs, because these interventions are designed to deliver tailored and timely support based on momentary context, vulnerability, and receptivity, rather than static content delivered uniformly across participants [16]. As a result, intervention groups often received more personalized prompts, summaries, feedback, adaptive logic, or supportive features than comparator groups, making performance bias difficult to avoid.

Meanwhile, non-controlled studies frequently showed weaknesses in selection procedures, as comparison groups were often absent and allocation methods lacked detailed justification. This limits confidence that observed effects can be attributed to the JITAIs being evaluated rather than to pre-existing participant differences or co-occurring influences. Again, this limitation may be especially important for JITAIs, because their effects are hypothesized to depend on timing, context, and adaptive delivery, making it necessary to isolate the contribution of specific intervention components [16]. Further, both randomized and non-controlled studies in our review demonstrated consistent weaknesses in outcome measurement reliability, participant retention, and statistical conclusion validity. Few studies provided sufficient study-specific evidence on measurement reliability, and loss to follow-up was often incompletely reported. Further, comprehensive details on statistical power and power assumptions were frequently absent or reported with caveats, consistent with the predominance of pilot samples across the included studies.

Within qualitative studies, the most consistent limitation was researcher positioning and reflexivity, which was rarely reported or addressed. This was reflected in limited reporting of whether the researcher’s cultural or theoretical orientation was declared. Further, the potential influence of the researcher on study participants, or vice versa, was rarely acknowledged. Health outcomes addressed by JITAIs for AYAs

Among JITAIs for AYAs, distal outcomes and their related proximal outcomes demonstrate some consistent priorities as well as emerging trends, with diverse conditions as the target of tailored support. We organize key outcomes targeted in JITAIs for AYAs into three domains: substance use, mental health, and physical health (Table 1). While substance use typically spans both mental and physical health, we present it separately here as it has been the most frequent and distinct focus of JITAIs for AYAs. Further, while we highlight some notable observed effects as well as inconsistencies, all results we report below should be interpreted with caution given the aforementioned methodological limitations and risks of bias detected across the evidence base.

**Table 1:**
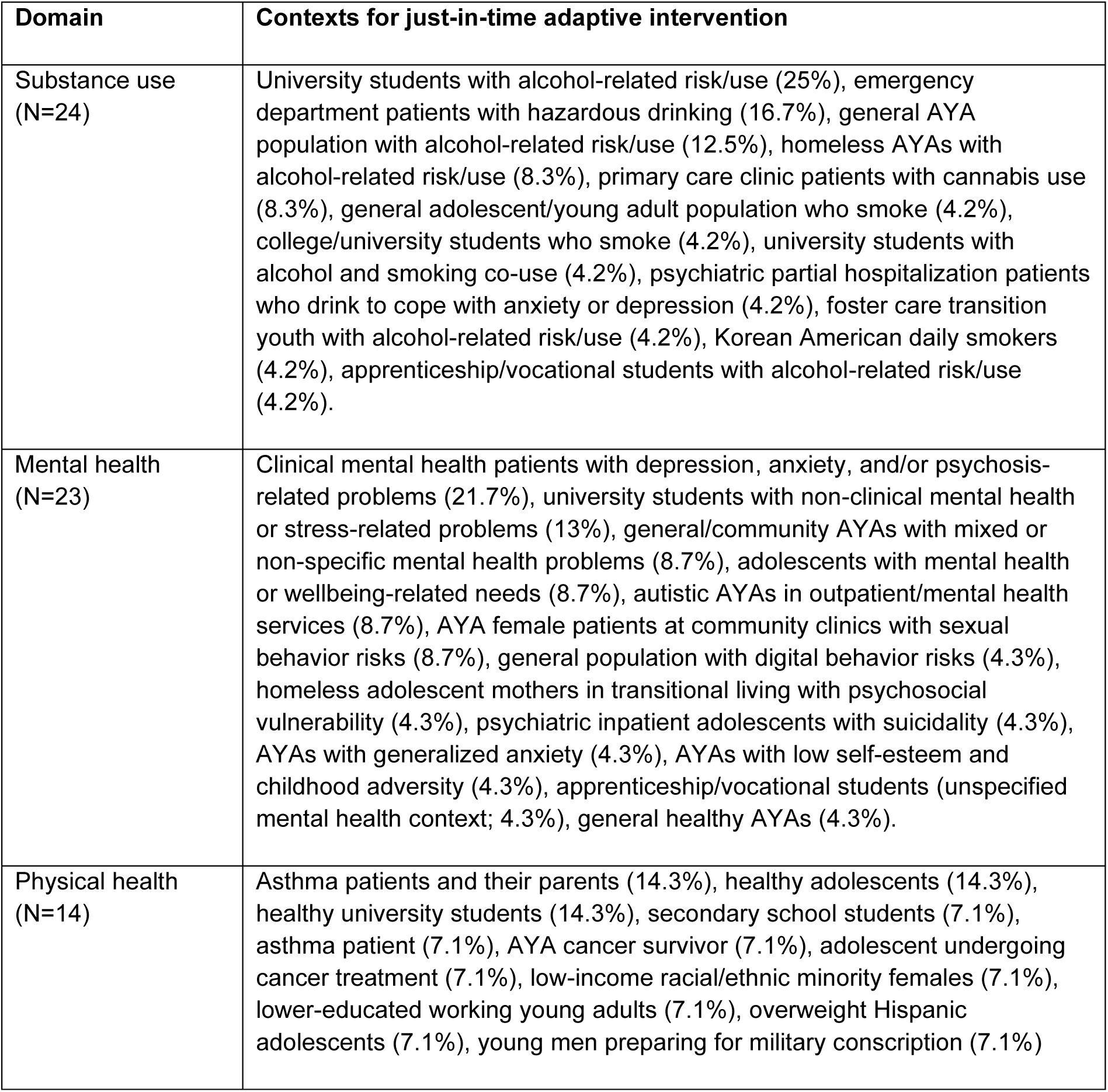
Contexts of JITAIs for AYA substance use, mental health, and physical health.

| Domain | Contexts for just-in-time adaptive intervention |
| --- | --- |
| Substance use<br>(N=24) | University students with alcohol-related risk/use (25%), emergency department patients with hazardous drinking (16.7%), general AYA population with alcohol-related risk/use (12.5%), homeless AYAs with alcohol-related risk/use (8.3%), primary care clinic patients with cannabis use (8.3%), general adolescent/young adult population who smoke (4.2%), college/university students who smoke (4.2%), university students with alcohol and smoking co-use (4.2%), psychiatric partial hospitalization patients who drink to cope with anxiety or depression (4.2%), foster care transition youth with alcohol-related risk/use (4.2%), Korean American daily smokers (4.2%), apprenticeship/vocational students with alcohol-related risk/use (4.2%). |
| Mental health<br>(N=23) | Clinical mental health patients with depression, anxiety, and/or psychosis-related problems (21.7%), university students with non-clinical mental health or stress-related problems (13%), general/community AYAs with mixed or non-specific mental health problems (8.7%), adolescents with mental health or wellbeing-related needs (8.7%), autistic AYAs in outpatient/mental health services (8.7%), AYA female patients at community clinics with sexual behavior risks (8.7%), general population with digital behavior risks (4.3%), homeless adolescent mothers in transitional living with psychosocial vulnerability (4.3%), psychiatric inpatient adolescents with suicidality (4.3%), AYAs with generalized anxiety (4.3%), AYAs with low self-esteem and childhood adversity (4.3%), apprenticeship/vocational students (unspecified mental health context; 4.3%), general healthy AYAs (4.3%). |
| Physical health<br>(N=14) | Asthma patients and their parents (14.3%), healthy adolescents (14.3%), healthy university students (14.3%), secondary school students (7.1%), asthma patient (7.1%), AYA cancer survivor (7.1%), adolescent undergoing cancer treatment (7.1%), low-income racial/ethnic minority females (7.1%), lower-educated working young adults (7.1%), overweight Hispanic adolescents (7.1%), young men preparing for military conscription (7.1%) |

### JITAIs addressing AYA substance use

Substance use reduction has been a consistently central focus of JITAIs for AYAs, with interventions (39.3%, N=24/61) targeting reduction in areas such as smoking cessation, alcohol moderation, and marijuana use [39–62]. The most common intervention contexts were for college or university students with alcohol-related risk or use (25%) and emergency department patients with hazardous drinking (16.7%). Novel population-specific pilot studies included interventions with homeless young adults as well as AYAs in foster care, respectively, with alcohol-related risks, as well as clinic patients with cannabis use [45,54,61].

JITAIs addressing substance use among AYAs typically emphasize regulating urges and reinforcing commitment to abstinence, often using momentary tailoring based on triggers and desire/urge state [39,45,49,51,55,61]. AYA JITAIs targeting smoking cessation demonstrated positive outcomes in several studies, including higher engagement linked to reduced and lower likelihood of smoking reported in one study [44]. Similarly, marijuana use reduction interventions also yielded encouraging results. Evidence includes higher abstinence at 6 weeks in an RCT (RR 2.20, 95% CI 1.79–2.70) [39]. Another pilot randomized trial found reduced odds of marijuana use after contextual or behavioral triggers (OR 0.54, 95% CI 0.31–0.95), alongside greater decline in momentary marijuana desire [45]. Additionally, interventions aimed at reducing drug use showed some promise, including in an RCT with AYAs experiencing homelessness with lower odds of reported drug use over time [49,61].

Alcohol-related outcomes were more mixed. Some studies reported reductions, including reduced heavy drinking days and drinks per drinking day. One emergency department sample found longer-term benefits, where interactive text messages reduced binge drinking and alcohol-related injury, with one third fewer binge drinking days at 9 months compared with controls (IRR 0.69, 95% CI 0.60-0.79) [47,63,64]. In contrast, a small but non-significant increase in consumption during heavy drinking occasions was reported elsewhere [46]. Other studies found most limited or non-significant between-group differences in alcohol outcomes [46,50]. Another intervention increased protective behavioral strategies even when drinking outcomes did not differ [48].

### JITAIs addressing AYA mental health

JITAIs with a specific focus on AYA mental health and well-being emerged in 2017 and have been steadily growing since (37.7%, N=23/61). These include interventions that target anxiety, depression, psychological distress, and suicide prevention among AYAs [65–81]. Further, JITAIs that address AYAs’ well-being more generally (as opposed clinical symptoms) focus on enhancing emotional resilience and coping strategies [82–87], including after experiencing cyber-victimization [88]. A development since 2024 includes 2 studies focused on promoting healthy digital behaviours, specifically to address the over-use of digital devices [89,90]. Intervention contexts were highly heterogeneous, with some JITAIs designed for clinical depression, anxiety, or psychosis-related problems (21.7%). Pilot studies included interventions conducted with autistic AYAs in outpatient services, female community clinic patients with sexual risk behaviors, and homeless adolescent mothers in transitional living programs [78,84,87]. Further, JITAIs for AYA mental health were supplements to in-person counseling or support group programs in several studies (39.1%), although the structure of these programs (e.g. individual versus group sessions, therapeutic framework) differed greatly.

Several AYA JITAIs targeting anxiety and depression symptoms demonstrated promising improvements across follow-up assessments, indicating potential effectiveness in alleviating specific clinical symptoms. Two RCTs reported promising mental health effects, with one trial observing significant reductions in depressive symptoms (β=-0.34, p=0.004), anxiety symptoms (β=-0.10, p=0.013), and perceived stress (β=-0.18, p=.035), while another found greater reductions in anxiety than in the control group (GAD-7: F(1,51)=9.06, p=0.004; d=0.61), with a nonsignificant trend for depression (PHQ-8: F(1,51)=3.21, p=0.08; d=0.50) [71,81]. However, less definitive results were observed in other studies. In one suicide prevention trial, no significant differences were found between intervention and control groups in suicide attempts or suicidal ideation, although effects were in the hypothesized direction [66,68]. Meanwhile, findings for general mental well-being outcomes were mixed: some interventions reported reduced perceived stress or improved self-esteem and confidence, whereas others found no significant effects on stress outcomes [73,81,82,84].

### JITAIs addressing AYA physical health

AYA physical health JITAIs (22.9%, N=14/61) often addressed obesity risk through the promotion of physical activity and healthy diets in distinct populations (e.g., racial minority groups) [91–100], although some studies have also focused on the management of chronic conditions, such as pain in cancer management as well as asthma [101–104]. Physical health strategies in these JITAIs often focused on incremental shifts, such as reducing sedentary behavior and promoting daily walking [94,95,98,99,92]. AYA JITAIs focused on nutrition encouraged immediate healthy choices to mitigate obesity risks [91,96,97,100]. Meanwhile, AYA JITAIs for chronic condition management promoted adherence behaviours (e.g., to medication) as well as pain management support, incorporating hybrid “clinical escalation” logic, such as nurse practitioner alerts triggered by diary thresholds and symptom ratings in pediatric oncology pain support, as well as caregiver and clinical staff alerts triggered by safety rules in an asthma action-plan app [101–103].

Physical activity promotion outcomes were variable. Modest increases in daily physical activity levels were observed, although these changes did not always extend to reductions in sedentary time [98,99]. For example, one study showed an initial decrease in sedentary minutes following intervention (t(17)=-1.79, p=0.04), though this effect attenuated over time and became non-significant by the second and third weeks [99]. Interventions targeting obesity-related risks also demonstrated limited effectiveness, with no significant changes in key indicators such as screen time and body mass index (BMI). However, some evidence indicated improvements in diet such as increased fruit and vegetable consumption and decreased sugar-sweetened beverage intake [91]. Finally, in a study specifically targeting cancer pain management, the intervention group reported fewer instances of moderate to severe pain compared to control (t(2145)=2.67, p=0.008, 95% CI 0.0036-0.0235), although there were no significant differences in average daily pain reduction [102].

### Theoretical frameworks informing intervention options

Table 2 presents a domain-level synthesis of the theoretical frameworks that informed (distinct) intervention designs, alongside the primary intervention options, tailoring variables, and decision points. Note that counts overlap as studies applied multiple components simultaneously. Across all JITAIs targeting AYA health, researchers commonly used tailored digital messages, which were delivered via text or app-based messaging tools, while leveraging diverse theoretical frameworks to inform intervention content.

**Table 2:** Summary of theoretical frameworks, intervention options, tailoring variables, and decision points in JITAIs for AYA substance use, mental health, and physical health.

| Domain | Substance use (N=24) | Mental health (N=23) | Physical health (N=14) |
| --- | --- | --- | --- |
| **Theoretical frameworks** | Motivational interviewing and related approaches (33.3%); cognitive-behavioral and related approaches (25%); transtheoretical model (16.7%); self-regulation (12.5%); information-motivation-behavioral skills model (8.3%); harm reduction (8.3%); not reported (12.5%). | Cognitive-behavioral and related approaches (34.8%); emotion regulation approaches (13%); motivational interviewing (13%), mindfulness-based approaches (13%), transdiagnostic approaches (13%), compassion-focused approaches (8.7%). | Self-regulation and control-based approaches (42.9%); Behavior Change Wheel (21.4%); positive psychology (14.3%); self-determination theory (14.3%); not reported (14.3%). |
| **Intervention options** | Psychoeducation modules (66.7%); supportive messages (62.5%); self-monitoring assessment (58.3%); goal-setting/ planning support (33.3%); interactive activities (33.3%); protective strategies/ coping | Coping and emotional support messages (65.2%); psychoeducation modules (65.2%); therapeutic skills practice (47.8%); self-monitoring check-ins (39.1%); interactive activities (30.4%); feedback/ progress summaries | Interactive activities (71.4%); psychoeducation modules (71.4%); suggestions/ feedback (71.4%); goal setting (57.1%); reminders (57.1%); self-monitoring/ tracking (50%). |
|  | support (33.3%); reminders (20.8%). | (26.1%); reminders (21.7%). |  |
| **Tailoring variables** | Self-reports of behavior (66.7%); motivation/ readiness (62.5%); triggers/ high-risk situations (45.8%); emotion/ affect (33.3%); goal attainment (33.3%); urges (29.2%); self-efficacy level (20.8%). One study collected electrodermal activity [53] | Self-reported emotion/ affect (65.2%); self-efficacy (21.7%); cognitive state (21.7%); behavior/ recent activity (34.8%); goals progress (17.4%). Passive collection included geolocation (17.4%), device usage (8.7%) and electrodermal activity [53] | Self-reported behavior (71.4%); clinical symptoms (35.7%); goals progress (50%); triggers/ high-risk situations (28.6%). Passive collection included sedentary time (28.6%), step count (28.6%), geolocation (21.4%), and food purchase history (N=1) [100] |
| **Decision points** | Daily (58.3%); event-triggered (50%); fixed scheduled time (37.5%); weekly (33.3%); on-demand/ user initiated (25%) | Daily (82.6%); fixed scheduled time (65.2%); on-demand/ user-initiated (39.1%); event-triggered (47.8%); weekly (30.4%) | Daily (71.4%); fixed scheduled time (50%); weekly (42.9%); event-triggered (35.7%); on-demand/ user-initiated (14.3%) |

In AYA JITAIs targeting substance use, motivational approaches (31% of substance use studies) was most frequently applied (33.3%), informing modules on self-reflection and dynamic goal-setting [43,45,47,51,53,54,58,60,62]. Cognitive-behavioral approaches were also frequently applied (25%), such as to inform coping prompts for managing substance cravings and related emotional distress that may trigger use [44,50,53,55,56]. Several JITAIs for AYA substance use also adopted the transtheoretical model (16.7%), which guided the tailoring of feedback specific to an individual’s stage of behavior change [40,54,57,60]. Among JITAIs focused on AYA mental health, most (34.7%) also leveraged cognitive-behavioral approaches to inform intervention content around cognitive restructuring and stress-related coping strategies [66,71,77,78,80,87,88]. JITAIs for AYA mental health also consisted of supportive messages based on emotion regulation [68,86,87], motivational interviewing [68,78,88], mindfulness-based strategies [72,79,87], in addition to transdiagnostic approaches [70,71,73], during heightened stress events (13% for each respective approach). Similarly, during individually challenging moments, compassion-focused JITAIs (8.7% within this domain) deliver content aimed at promoting emotional resilience and self-kindness [65,67,70].

For JITAIs promoting AYA physical health, self-regulation and control-based approaches were most frequently adopted (42.9%) to tailor motivational prompts that emphasize autonomy and self-efficacy to encourage sustained health behaviours [91,93,97–99]. 21.4% of JITAIs for AYA physical health also adopted the Behaviour Change Wheel to guide structured prompts and reinforcement strategies, such as reminders to reduce sedentary behavior and promote physical activity, including supplementation with gamified reward systems [96,97,99]. Among JITAIs focused specifically on managing physical symptoms of chronic health conditions, frameworks such as self-regulation theory were used to structure feedback mechanisms to address adherence barriers, such as for asthma management [104].

There is an overall trend of basing adaptive content on theories related to promoting AYAs’ levels of motivation and self-efficacy, states of cognition and behavior change, although the specific application of these principles varies by context. For example, cognitive-behavioral approaches can be broadly applied to aid in managing cravings in substance use in addition to cognitive restructuring in mental health, while also supporting adherence behaviors in chronic pain management. Behavioral intent and goal-setting also inform intervention content, with JITAIs commonly adapting their support based on AYA-defined goals and progress [64,91,93,98,99,105]. This significant variability underscores the further need to understand how the adaptive selection of these dynamic interventions, such as timing, content, and delivery, align with ongoing AYA contexts, which is the focus of the next section.

### Tailoring variables, decision rules, and decision points

Tailoring mechanisms within AYA JITAIs are primarily informed by behavioral, psychological, and other contextual data (Table 2). Further, the tailoring variables were aligned with the proximal and distal outcomes of the interventions. For example, behavioral data, such as self-reported nutritional consumption and physical activity levels was used to prompt personalized feedback and inform the content of messages [93,97,98]. Meanwhile, psychological variables, such as fluctuations in mood and self-efficacy, informed JITAIs aimed at improving AYAs’ emotional regulation and reducing negative affect [66,68,70,71,78]. Across these approaches, contextual awareness is emphasized, with JITAIs tailoring content based on individual and environmental factors.

Among AYA JITAIs addressing substance use and mental health, most tailoring variables rely on self-reported data, such as mood, stress, and alcohol consumption. Substance use interventions predominantly tailored content to self-reports of substance use (e.g., frequency, quantity, cravings, triggers; 66.7%); additional tailoring drew upon motivational states (62.5%) as well as high-risk social or situational contexts (e.g., location, time of week, social setting; 45.8%). One study collected electrodermal activity to determine high-risk stress states requiring intervention to prevent drinking [53]. Mental health-focused JITAIs tailored primarily to self-reported affective states (65.2%), such as mood, stress, and negative thinking. Some studies also tailored support based on self-reported self-efficacy and coping needs (21.7% respectively). Passive collection included geolocation (17.4%), device usage (8.7%) and one study leveraging electrodermal activity to determine states of stress [53].

In contrast to JITAIs for AYA substance use or mental health, over half of JITAIs for AYA physical health tailored intervention delivery based on passive activity data. These include tailoring based on step counts (28.6%) and sedentary time (28.6%), including via sensing data using Bluetooth geofencing (21.4%). This reflects a trend toward real-time, event-triggered decision points in JITAIs, characterized by interventions that are activated based on immediate sensor data, such as detecting physical inactivity [94,95,99]. Notwithstanding, self-reported data was also frequently leveraged in JITAIs for AYA physical health, including for monitoring behavior (71.4%), chronic disease symptoms (35.7%), and goals progress (50%).

In terms of decision points, JITAI delivery is characterized by the use of daily or multiple times daily surveys to collect these tailoring variables [91,98,101–104]. These decision points enabled AYAs to prompt immediate interventions adapted based on the self-report. Across all AYA JITAIs, these mechanisms can also be broadly characterized as following threshold-based triggers, where responses are activated if pre-defined conditions are met, such as reaching a specific stress level or failing to meet activity goals [43,53,55,56,65,66,68,70,72,81,99,101,103]. This approach allowed for adaptive interventions to be activated after consistent patterns of inactivity, poor mental health, or unmet goals. Such mechanisms were further enabled through the incorporation of high-frequency, scheduled prompts, including multiple daily check-ins or morning and evening notifications [48,50,51,69,72,81,98,101,104].

Some studies also allowed for AYA-initiated check-ins, to promote flexibility in need-based and real-time support [40,53,59,69,70,76,88]. Meanwhile, in studies which employed behavioral monitoring, interventions were characterized by fixed-schedule decision points, which were based on predefined intervals [50,54,66,81,83,91,98,102,104]. Studies also tailored intervention schedules based on participant-defined or behaviorally relevant times, such as following peak drinking days [43,47,50]. Time of day and spatial cues have also been leveraged to refine the timing and delivery of interventions, with methods including geofencing used to automatically identify high-risk locations and detecting sedentary behaviours to prompt tailored notifications [46,50,75,76,99].

Since 2024, JITAIs for AYAs have leveraged intelligence to enhance the adaptability of interventions. One study developed a JITAI to prevent smartphone overuse, introducing a friction-based intervention to disrupt access to blacklisted apps when overuse was detected. The authors employed a machine learning model which was updated daily with passive sensing data such as app usage, activity, social context, location, and temporal patterns to predict overuse, triggering interventions upon app launches and task-switching moments [89]. Another recent study leveraged large language models (LLMs) for dynamic adaptability, also with the goal of reducing problematic smartphone use [90]. Similarly, when AYA users opened blacklisted apps, the LLM generated persuasive messages using four communication strategies of understanding, comforting, evoking, and scaffolding. LLM messages were also aligned with tailoring variables, which included real-time usage, context, mental states, and user-defined goals related to app use.

### Ethical considerations in JITAIs for AYAs

A critical gap emerges in the lack of detailed reporting on AYA engagement across studies, limiting formal analysis with the tailoring mechanisms identified in prior sections. AYA engagement in the development of JITAI components demonstrates considerable variation in both depth and rigor. Certain studies integrated current participants directly into the design process [58,60,97,104], while others consulted AYAs with demographic profiles similar to the intended intervention audience [39,79,85]. Such engagement is frequently framed as a strategy to enhance intervention relevance and acceptability by involving adolescents in consulting on the relevance of content. However, the extent of AYA participation in these studies is also inconsistently reported, particularly within RCTs, which often prioritize the evaluation of pre-developed interventions over documentation of co-design processes. In contrast, qualitative studies focused on describing processes of adolescent engagement in JITAI component development explicitly.

While unreported involvement in these interventions is also possible, the absence of such documentation limits our current ability to assess their impact on JITAI tailoring mechanisms. Moreover, among studies claiming AYA involvement, the depth of engagement was frequently superficial. It was unclear if AYA had substantive influence over core design decisions regarding decision rules, except for among the small sample of qualitative studies. For example, one study engaged AYAs exiting foster care in co-designing a JITAI for substance use reduction [54]. Through focus groups with these AYAs, participants provided iterative feedback on intervention structure, language, message frequency, and content relevance, which led to key modifications in JITAI content, including adjustments to message tone, content specificity, and delivery timing. Similarly, one study involved Dutch lower-educated AYAs in the iterative development of JITAI content, including informing modifications in text clarity, visual elements, navigation, and notification settings [62]. Another study involved ethnic minority adolescent females in developing a Build Your Own Theme Song app [77]. Adolescent females contributed to JITAI design by creating personal theme songs, through rewriting and recording lyrics based on their experiences. Subsequently, the app would prompt adolescents to engage with their theme song during moments of negative thinking, providing a personalized coping mechanism based on their own input.

Ethical considerations were rarely addressed in detail for specific intervention components or data collection mechanisms. A minority described explicit strategies that were primarily limited to study design, such as providing devices or data plans [44,53,87], using data encryption [101], or advising participants on secure message deletion [66]. However, reporting on the ethical implications of tailoring mechanisms were notably absent. No studies discussed mechanisms for data sovereignty or the granularity of consent regarding how distinct personal data might inform algorithmic and intervention decision-making. Despite the increasing use of passive sensing and machine learning [89], there was a lack of transparency regarding how these automated decisions were explained to adolescent users. This obscures potential risks related to privacy and autonomy, highlighting an urgent need for standardized ethical reporting guidelines in AYA digital health.

## DISCUSSION

This systematic review identified key patterns in how JITAIs for AYAs utilize dynamic tailoring mechanisms, decision rules, and adaptive data sources to personalize health interventions to improve substance use, mental health, and physical health (including chronic disease) outcomes. Most JITAIs relied on self-reported data, with growing trends towards integrating passive and sensor-based data, such as geofencing, app usage, and physiological monitoring [66,76,87,89,91]. Decision rules were frequently based on threshold triggers or scheduled intervals, though emerging studies have begun to explore LLMs and machine learning models for real-time, adaptive interventions [89,90].

The dominance of self-report of behavioral data in JITAIs also suggests limited consideration of broader contextual and social determinants of health in their tailoring mechanisms, even when studies were piloted specifically with underserved communities [49,61,62,77,87,91]. Moreover, while our risk of bias appraisal assessed participant retention across studies, JITAI-specific data missingness also includes ecological momentary assessment non-response, sensor data loss, or selective absence of data at clinically important moments [106,107], which likely also influenced outcome measurement quality as well as intervention delivery. It is crucial for future work to investigate the incorporation of both social and missingness factors in JITAI tailoring to enhance contextual relevance and precisely address barriers to equitable health support among diverse individuals [27,108,109].

Moreover, JITAI mechanisms were highly heterogeneous and confidence in the evidence base was constrained by inconsistent reporting and heterogeneous study designs. Further, while specific outcomes have yielded positive effects, most outcomes exhibit more variable or limited findings [44,46,50,82,92,93]. Taken together, these patterns suggest the need for further investigation into the methodological determinants of intervention success, particularly regarding how JITAI and study design influence observed effectiveness.

### Limitations of existing JITAI study designs

It is crucial to interpret current observations of effectiveness in this review, including limited effects, with significant caution. The majority of current evidence for AYA JITAIs comes from small, pilot uncontrolled experiments. Comparison groups were often absent or weak and attrition or response burden was common. Several authors also explicitly noted that observed improvements could not be disentangled from self-monitoring effects, baseline reporting, treatment as usual, in-person counseling, or other concurrent influences [51,56,65]. Meanwhile, while a substantial number of studies were randomized, the risk of bias assessment highlighted recurrent challenges such as limited blinding, unclear allocation concealment, and non-identical intervention administration. Heterogeneity in comparators also reduce confidence that the current observed effects can be attributed solely to the JITAIs being tested [110].

Given these limitations, the literature may particularly benefit from not further RCTs, but more optimization-oriented trials, particularly microrandomized trials (MRTs) which can evaluate which intervention components work, at what moments, and for what specific AYA daily life contexts. MRTs, in contrast to conventional RCTs, randomize intervention options at multiple and repeated decision points to help precisely determine the effectiveness of individual adaptive components across diverse individual contexts [111]. Such an approach can enable “a holistic comprehension of intervention dynamics over various timescales” [108]. We posit that even randomized JITAI study designs may be unable to sufficiently detect within-person variability or isolate momentary effects over time, and therefore may potentially underestimate or construe interpretations of the effectiveness of JITAIs. To deliver precise JITAIs to AYAs, it may be important to assess diverse contextual factors which may influence the effectiveness of different adaptive components, as this can help understand “when and in what contexts a particular intervention option should be delivered” – a key differentiator of JITAIs from other digital health interventions [111]. In sum, MRTs can crucially more precisely enable the measurement of the specific tailoring variables, decision points, and intervention options that are optimal for different individuals and health outcomes [108]. MRTs can also help to estimate proximal effects even under variable engagement and contextual missingness.

Further, for qualitative studies, the risk of bias assessment highlighted that many co-authors did not sufficiently consider researcher positioning and reflexivity [112]. AYA participation in JITAI development was often limited to feedback on usability or content refinement rather than shared involvement in intervention design. We also observed inconsistent considerations of privacy, consent, and ethical safeguards in AYA JITAIs. These findings highlight critical gaps in ensuring that JITAIs align meaningfully with AYA contexts and needs alongside critical ethical standards [113].

### JITAIs for AYAs: preliminary recommendations for future directions

To address limitations of existing JITAI study designs, we sought interpretive feedback from AYAs regarding our current evidence base (see Methods section for process). Their contributions here provide a complementary perspective on the evidence, highlighting gaps and priorities for future JITAI research from their perspectives as target users.

Despite current JITAIs incorporating frequent rule-based adaptations, AYAs we consulted showed a preference for simplicity and user-controlled customization of intervention options. They highlighted how current JITAI tailoring mechanisms are overly complex or opaque. They also mentioned the lack of modular designs, whereby users can select which features they wish to engage with to enhance contextual relevance. Ongoing notifications, which characterize many of the JITAIs in this review [39,54,69,81,91,98,99], were criticized for potentially contributing to notification fatigue which may limit engagement over time. For example, our AYA consultants discussed how excessive prompts can lead to desensitization, frustration, and eventual disregard of JITAIs. Subtle, passive engagement strategies, such as visual cues or widgets which update in the background of devices and can be checked flexibly, were highlighted as a potential alternative [114]. This raises the possibility that high-frequency prompting may undermine engagement or perceived usefulness, particularly among AYAs already experiencing digital overload and other digital determinants of poor health [115,116]. Our AYA consultants also raised concerns about data privacy and lack of control over what information is collected, advocating for opt-in tailoring mechanisms that allow users to choose which data they are comfortable sharing [117].

This feedback from our AYA consultants regarding our current evidence base highlight a potential tension within current JITAI development and trends towards passive sensing and multimodal integration with artificial intelligence [118]. In an effort to enhance engagement and precision, these mechanisms may also risk reducing engagement or acceptability for some AYAs [119]. Our AYA co-authors described this phenomenon as an “engagement paradox” – a concept emerging from the divergence between the high-frequency adaptability reported in the literature and the desire for autonomy expressed by our AYA consultants. This may indicate that the success of adaptive interventions depends not only on the incorporation of multiple levels of tailoring, but also on how well tailoring mechanisms align overall with AYAs’ ongoing preferences and need for autonomy [120,121]. Despite automation’s role in enhancing adaptation, our AYA consultants suggested that excessive automation may reduce AYAs engagement if the tailoring mechanisms lack transparency and explainability. As such, adaptive systems where AYAs can define their preferred tailoring mechanisms may be a worthwhile direction for future precision health interventions. Transparent data practices, which can promote participant control over tailoring mechanisms, may also be essential to privacy considerations as well as intervention relevance [122,123].

Notwithstanding, both the AYAs we consulted and engaged in this review as co-authors were limited in number and may not represent other AYAs’ perspectives. Substantial further participatory research is necessary to improve the precision and relevance of JITAIs for diverse AYA lived experiences [108]. To provide a foundation for such work, we present preliminary recommendations for future directions to explore in investigations of AYA JITAIs.

### Investigate mechanisms of meaningful engagement in real-world contexts

Understanding the mechanisms by which JITAIs influence behavioral outcomes is critical for advancing the field [16,108]. The review highlighted uncertainties regarding how adaptive mechanisms operate and influence engagement and outcomes, alongside key limitations of existing JITAI study designs. To address these challenges, future research can employ optimization trial designs [111], such as micro-randomized or sequential multiple assignment trials to systematically test how specific tailoring variables, such as timing, content format, and data type, as well as theoretical frameworks may impact engagement and behavioral outcomes. This can enable a more precise understanding of how different intervention components and decision rules perform, and help clarify the tailoring mechanisms which are most effective for specific AYA contexts [108].

Further, more meaningful co-design in creating relevant, effective interventions is a crucial next step [124]. The review revealed limited AYA involvement in the design of many JITAIs, resulting in interventions that may not fully align with users’ lived experiences or preferences [109]. Insights from our consultation with AYAs reinforced this, highlighting the value of relevance and understandability in design. There is a need to embed precision health designs within real-life contexts to ensure interventions are adaptable and impactful for ongoing AYA health needs [108,109,125]. Participatory workshops can facilitate meaningful input, enabling AYAs to contribute ideas about preferred tailoring mechanisms. For example, studies can engage adolescent participants in informing the proximal outcomes, intervention options, tailoring variables, decision points, as well as overall user experience, of JITAIs [54]. Integration of participatory design components to additionally inform as well as interpret individual outcomes in ongoing MRTs would enable more comprehensive understanding of what works well, for whom, and why [126]. Such approaches would help acknowledge AYAs as experts by experience, resulting in interventions that account for and are more grounded in real-life contexts [127]. In this manner, AYAs can directly participate in shaping mechanisms in precision health interventions.

### Continuous adaptation directed by adolescents

Adaptation is a cornerstone of effective JITAI design [16], yet the review findings identified notable gaps and inconsistencies in how such adaptation is actually implemented. Further, our AYA consultants, in reviewing our evidence base, expressed alternative preferences for systems that allowed for greater control and customization rather than opaque, automated adaptations. To address these concerns, future JITAIs can incorporate modular, user-directed adaptation features that empower adolescents to adjust the level and type of adaptation as their preferences and needs evolve [122,128]. For instance, intervention component dashboards could be developed, allowing AYAs to select their preferred notification frequencies, data sources for tailoring, and engagement formats, such as text, video, or visual cues [62,129–131]. Such affordance of ongoing participatory adaptation may help promote AYA agency while fostering enhanced trust and relevance, ensuring that JITAI mechanisms are both transparent and AYA-centered. However, we acknowledge that such open customization introduces heterogeneity in intervention exposure, potentially complicating the assessment of internal validity [132]. To address this, we again recommend that future research utilize optimization trial designs, such as MRTs, which are uniquely capable of handling within-person variability [133]. By treating user-defined preferences as dynamic tailoring variables within the statistical model, researchers may be able to better understand the mechanism of such autonomy, separate from the effects of the content itself.

### Minimizing engagement fatigue while promoting more meaningful engagement

The systematic review identified inconsistent outcomes and effects among current JITAIs for AYAs, which may be in part attributable to the methodological heterogeneity or lack of consistent theoretical grounding across studies. Further, based on the interpretive feedback from AYAs we consulted, we posit that low user engagement may also be driven by “notification fatigue” [134] – a phenomenon they identified as a critical barrier to sustained use. While the empirical studies in our review rarely measured fatigue explicitly, AYAs we consulted criticized frequent and intrusive notifications for contributing to disengagement. To address these challenges, future intervention development might explore alternative, less intrusive engagement strategies [135]. For example, AYA JITAIs might explore replacing frequent text notifications with visual nudges or contextual prompts [62]. Interventions could also explore integrating prompts into commonly used platforms, such as social media feeds, to ensure engagement occurs within familiar and comfortable digital spaces to enhance relevance to AYAs [136,137].

### Strengthening ethical standards for transparent personalization and digital self-determination

The review identified significant gaps in ethical reporting, with most studies neglecting to mention ethical considerations in JITAI design mechanisms. Such considerations are particularly critical given JITAIs which rely on sensitive data collection and passive monitoring [138]. Such an approach also aligns with principles of digital self-determination, which emphasize the reduction of information asymmetries by ensuring adolescents have control over their data and tailoring mechanisms [130]. Our AYA consultants also expressed discomfort with unclear or opaque data practices of current JITAIs, and emphasized a strong preference for opt-in mechanisms that allowed them to control the type and extent of data they shared. In this context, a key consideration in future JITAI development may include the need to promote AYAs’ individual agency over their digital data and enable participatory tailoring mechanisms [122,139].

To address these concerns, dynamic consent processes within ongoing JITAI tailoring mechanisms may be one promising approach. Dynamic consent refers to when individuals are continuously informed and able to manage their data-sharing preferences over time [140]. Such a practice can enhance data sovereignty, of the need for individuals to “have control over the use of their shared data,” without which JITAI utilization may be significantly hindered [140]. These processes can provide clear, accessible information about what data is collected, how it is used, and the purpose of each data point in tailoring mechanisms. AYAs can be further be empowered to opt-in or opt-out of specific data types and be given the opportunity to modify data points over time. This can be operationalized through user-friendly privacy dashboards which enable real-time adjustment of intervention tailoring settings [140,141].

Dynamic consent in AYA digital health remains very limited. Recent studies highlight the approach’s alignment with technology users’ desire for transparency and control [140]. Studies have also shown how dynamic consent preferences are highly individual based on demographic and social contexts [131], making the approach particularly salient for application in precision adolescent and digital health contexts. Based on our AYA consultation, we posit that increased emphasis on self-determination and sovereignty in JITAI design may help improve their alignment and relevance with adolescent health needs and expectations.

### Strengths and limitations of the review

This review is the first to focus on JITAIs across a wide range of health conditions within a specific developmental period, highlighting the unique needs and intervention contexts for adolescents, who face distinct physical, mental, and social health challenges that differ from those of children or adults. A key strength of this review is the meaningful involvement of AYAs in screening, data extraction, and critical interpretation, which enhances the relevance and actionability of the findings and future recommendations. The current synthesis of common theoretical frameworks and tailoring mechanisms also represents an important first step towards a more coherent body of evidence for the application of JITAIs in AYA health. However, due to substantial heterogeneity in intervention designs, high risk of bias (particularly with outcome reliability and statistical validity), and inconsistent outcome effects observed, meta-analysis was not feasible. Additionally, despite a broad search strategy, some relevant studies may have been inadvertently missed, as most included interventions did not explicitly describe their mechanisms as just-in-time or adaptive.

## CONCLUSION

This review provides the first comprehensive synthesis of how JITAI mechanisms are being designed and studied for AYAs across multiple health domains, while also integrating AYA perspectives into the interpretation of findings and future directions. Beyond showing where JITAIs are being applied, it highlights important limitations in the current evidence base, including heavy reliance on self-reported behavioral data and limited attention to broader social determinants in tailoring mechanisms. The review also highlights substantial methodological heterogeneity among studies evaluating similar outcomes alongside insufficient ethical transparency, including participatory engagement to ensure intervention designs are grounded in real-life AYA contexts. Our findings suggest that progress in AYA precision health may depend not only on developing more adaptive interventions, but on measuring and designing them in ways that are more transparent and participatory. Limitations of traditional study designs, including measurement of JITAI administration in randomized controlled trials, point to the need for more robust designs such as MRTs as a key methodological next step for identifying which adaptive components work, at what moments, and in which daily life contexts. Based on AYA co-author and consultant input, we further outline preliminary recommendations for future research to help ensure JITAIs better align precision with AYA agency, including through co-designed tailoring mechanisms, reduced notification burden, and clearer control over personal data and automated decision-making.

## ACKNOWLEDGEMENTS

We thank Stella Goeschl, Anastasia Kluge, and Benedetta Vallese for their valuable input which informed the development of the manuscript. We also thank Dirk Jan Ligtenbelt, who provided guidance on the search strategy. Parts of this work were presented at the Karolinska Institutet – United Nations Children’s Fund Joint Conference on Global Child and Adolescent Mental Health 2024, the 19th European Association for Research on Adolescence Conference, and the Social Data Science research group seminar at the Alan Turing Institute; we thank attendees for their thoughtful reception and feedback on earlier versions of this work.

## DATA AVAILABILITY

All data extracted and reported in this review are available as supplementary material.

## AUTHOR CONTRIBUTIONS

KWG: review guarantor, conceptualization, project administration, methodology, investigation, formal analysis, writing – original draft, writing – review and editing. MA: investigation, formal analysis, writing – original draft, writing – review and editing. SG, IK, CA, VC, RA, CI, ET: investigation, writing – review and editing. CRS, AV, EvR: methodology, writing – review and editing. LK: funding acquisition, methodology, writing – review and editing. MdR: funding acquisition, supervision, writing – review and editing. CAF: funding acquisition, supervision, project administration, methodology, writing – review and editing. KWG used ChatGPT solely for limited editorial support during final manuscript preparation, specifically grammar correction and table formatting. ChatGPT was not used for review design, data analysis, interpretation, ideation, reference generation, or any substantive writing. No AI-generated content were included in the final manuscript. All outputs were reviewed and verified by the authors, who remain fully accountable for all aspects of the manuscript.

## ABBREVIATIONS

AYA: Adolescent and young adult
JITAI: just-in-time-adaptive intervention
RCTs: randomized controlled trials
MRTs: microrandomized trials

